# Amyloid and Tau PET positive cognitively unimpaired individuals: Destined to decline?

**DOI:** 10.1101/2022.05.23.22275241

**Authors:** Rik Ossenkoppele, Alexa Pichet Binette, Colin Groot, Ruben Smith, Olof Strandberg, Sebastian Palmqvist, Erik Stomrud, Pontus Tideman, Tomas Ohlsson, Jonas Jögi, Keith Johnson, Reisa Sperling, Vincent Dore, Colin L. Masters, Christopher Rowe, Denise Visser, Bart N.M. van Berckel, Wiesje M. van der Flier, Suzanne Baker, William J. Jagust, Heather J. Wiste, Ronald C. Petersen, Clifford R. Jack, Oskar Hansson

## Abstract

A major unanswered question in the dementia field is whether cognitively unimpaired individuals who harbor both Alzheimer’s disease (AD) neuropathological hallmarks (i.e., amyloid-β plaques and tau neurofibrillary tangles) can preserve their cognition over time or are destined to decline. In this large multi-center amyloid and tau PET study (n=1325), we examined the risk for future progression to mild cognitive impairment and the rate of cognitive decline over time among cognitively unimpaired individuals who were amyloid PET-positive (A+) and tau PET positive (T+) in the medial temporal lobe (A+T_MTL_+) and/or in the neocortex (A+T_NEO_+) and compared them with A+T- and A-T-groups. Cox proportional hazard models showed a substantially increased risk for progression to mild cognitive impairment in the A+T_NEO_+ (Hazard ratio [HR]=19.2[95% confidence interval: 10.9-33.7]), A+T_MTL_+ (HR=14.6[8.1-26.4) and A+T-(HR=2.4[1.4-4.3]) groups vs the A-T-(reference) group. Linear mixed effect models indicated that the A+T_NEO_+ (β=-0.056±0.005, T=-11.55, p<0.001), A+T_MTL_+ (β=-0.024±0.005, T=-4.72, p<0.001) and A+T-(β=-0.008±0.002, T=-3.46, p<0.001) groups showed significantly faster longitudinal global cognitive decline compared to the A-T-(reference) group (all p<0.001). Evidence of advanced AD pathological changes provided by amyloid and tau PET is strongly associated with short-term (i.e., 3-5 years) cognitive decline in cognitively unimpaired individuals and is therefore of high clinical relevance.

## INTRODUCTION

Alzheimer’s disease (AD) is neuropathologically characterized by the presence of amyloid-β (Aβ) plaques and tau neurofibrillary tangles. Although the two most well-established diagnostic criteria for AD both acknowledge the importance of Aβ and tau pathology in AD pathogenesis, an important distinction is that the National Institute on Aging and Alzheimer’s Association (NIA-AA) criteria^1^ define AD by its biological features (i.e., the presence of Aβ and tau pathology) irrespective of the clinical syndrome, while the International Working Group (IWG) criteria^2^ require the presence of objective cognitive impairment (mild cognitive impairment [MCI] or dementia) in conjunction with positive AD biomarkers. Consequently, there is fundamental disagreement between the criteria about the nomenclature for cognitively unimpaired individuals who harbor one or both AD hallmark neuropathological features. For example, a cognitively unimpaired individual with positive Aβ (β+) and tau (T+) biomarkers is classified as “preclinical AD” by the NIA-AA criteria^1^, while the IWG criteria would label such an individual “at risk for progression to AD” (**Fig.1a**).

**Fig 1.**
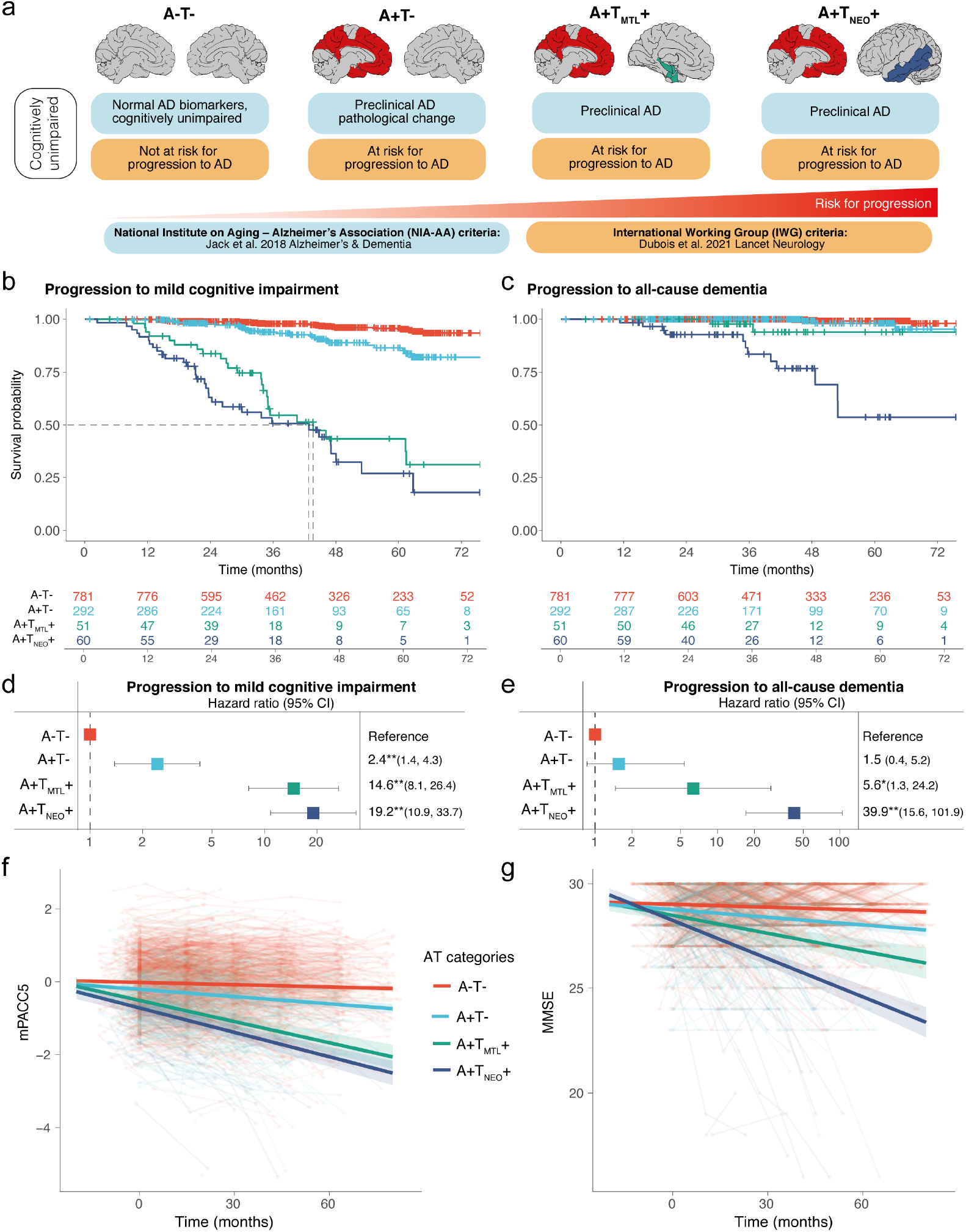
Progression to MCI or all-cause dementia and cognitive decline in the different AT categories **a**, Differences in nomenclature of cognitively unimpaired individuals with (+) or without (-) *in vivo* biomarker evidence of amyloid-β (A) and tau (T) pathology in the NIA-AA vs IWG criteria for Alzheimer’s disease. Note that for the IWG criteria the presumed “risk for progression” level rises when both A and T biomarkers are positive. **b**, Survival curves in relation to progression to MCI in the different AT categories, with the table of total number of participants available at each time point. The dashed indicates the time point at which 50% of a group had progressed to MCI. **c**, Survival curves in relation to progression to all-cause dementia in the different AT categories, with the table of total number of participants available at each time point. **d, e**, Forest plots showing the hazard ratios and 95% confidence intervals derived from the survival analyses shown in b and c, from Cox regression models including age, sex, education, and cohort as covariates. **f**, Cognitive trajectories of mPACC5 scores over time in the different AT categories. **g**, Cognitive trajectories of MMSE scores over time in the different AT categories. The average regression line for each group was plotted from linear mixed effect models including age, sex, education, and cohort as covariates. Data are anchored to the tau-PET visit (Time 0), and cognitive data up to 1 year prior to PET was included. *p<0.01, **p<0.001.

Aside from philosophical differences (i.e., according to the IWG criteria the term AD should be restricted to symptomatic individuals), the discrepancy between the NIA-AA and IWG can be explained by at least two factors. First, according to the IWG-criteria, currently available Aβ and tau biomarkers show “low predictive accuracy” for development of cognitive symptoms. For Aβ biomarkers alone this may indeed be the case^3,4^, although PET studies with long follow-up durations (∼10 years) indicate significant Aβ-associated risk for cognitive decline^5^, dementia^6^ and death^6^. For tau biomarkers, however, the clinico-pathological correlates are much stronger than for Aβ biomarkers, even in asymptomatic individuals.^7,8^ Second, most prognostic studies to date are based on cerebrospinal fluid (CSF) biomarkers of soluble phospho-tau levels, which is an early marker of AD pathology. In contrast, the more recently introduced technique tau-PET measures more advanced pathological changes as it captures insoluble tau aggregates.^9^ Although tau-PET positivity in the neocortex is relatively rare among cognitively unimpaired individuals (∼5-10%^10-12^), its presence is strongly associated with worse cross-sectional and longitudinal cognitive outcomes.^13,14^ However, large-scale longitudinal studies with amyloid-PET and tau-PET as predictors of clinical progression among cognitively unimpaired individuals are lacking.

Therefore, the aim of the current multi-center study was to examine short-term (i.e., 3-5 years) clinical progression to mild cognitive impairment (MCI) or dementia and to assess cognitive decline in cognitively unimpaired individuals with different Aβ (A) and tau (T) biomarker profiles as defined by PET. We divided A+T+ individuals into medial temporal lobe only (A+T_MTL_+) and neocortical (A+T_NEO_+) T+ groups to additionally investigate the impact of more advanced tau pathological changes on clinical progression.

## RESULTS

### Participants

We included 1325 cognitively unimpaired participants from seven cohorts, of whom 843 (63.6%) were A-T-, 328 (24.8%) A+T-, 55 (4.2%) A+T_MTL_+ and 65 (4.9%) A+T_NEO_+ (see **Table-1** and **Extended-Data Table-1** for a breakdown by cohort). All biomarker-positive groups were older and had lower baseline MMSE scores compared to the A-T-group (all p<0.001). There were no sex differences between groups. The average follow-up duration was 41.8±18.9 months. The A-T+ group was considerably smaller than the other groups (n=34, **Extended-Data Table-2**), hence their results are only reported in **Extended-Data Fig.1**.

**Table 1.** Participant characteristics

|  | A-T- | A+T- | A+T<sub>MTL</sub>+ | A+T<sub>NEO</sub>+ | P-value |
| --- | --- | --- | --- | --- | --- |
| N | 843 | 328 | 55 | 65 |  |
| Age, years | 68.6±9.6 | 75.5±8.2 | 75.6±6.6 | 76.4±6.8 | <0.001<sup>a</sup> |
| Sex, n (%) male | 424 (50.3) | 164 (50.0) | 24 (43.6) | 31 (47.7) | 0.822 |
| Education, years | 14.7±3.1 | 14.6±3.1 | 13.7±3.8 | 13.9±3.5 | 0.02<sup>b</sup> |
| Follow-up duration, months | 43.0±18.9 | 40.1±17.6 | 39.0±16.5 | 36.4±14.9 | 0.004<sup>c</sup> |
| Follow-up visits, number | 4.1±1.5 | 4.1±1.4 | 4.0±1.3 | 3.6±1.2 | 0.07 |
| MMSE, baseline score | 29.0±1.0 | 28.7±1.3 | 28.3±1.5 | 28.2±1.3 | <0.001<sup>d</sup> |

A = Amyloid; MMSE = Mini-Mental State Examination; MTL = Medial temporal lobe; NEO = Neocortical; T = Tau.

### Clinical progression to MCI

During clinical follow-up, 26/781 (3.3%) of A-T-, 26/292 (8.9%) of A+T-, 25/51 (49.0%) of A+T_MTL_+ and 32/60 (53.3%) of A+T_NEO_+ participants progressed to MCI or dementia. Cox proportional hazard models, adjusted for age, sex, education and cohort, showed an increased risk for future progression to MCI in the A+T_NEO_+ (Hazard ratio [HR]=19.2[95% confidence interval: 10.9-33.7], p<0.001), A+T_MTL_+ (HR=14.6[8.1-26.4], p<0.001) and A+T-(HR=2.4[1.4-4.3], p=0.002) groups compared to the A-T-(reference) group (**Fig.1b-d)**. Pairwise log-rank tests showed that the A+T_MTL_+ and A+T_NEO_+ groups (both p<0.001) had steeper survival curves compared to the A+T-group, while the A+T_MTL_+ and A+T_NEO_+ groups did not differ from each other (p=0.19). Fifty percent of the A+T_NEO_+ and A+T_MTL_+ groups had progressed to MCI after 42.8 and 43.6 months, respectively.

### Clinical progression to all-cause dementia

During clinical follow-up, 21 participants progressed to all-cause dementia; 4/781 (0.5%) in A-T-, 3/292 (1.0%) in A+T-, 2/51 (3.9%) in A+T_MTL_+ and 12/60 (20%) in A+T_NEO_+ (**Fig.1c-e**). Of those, 14 progressed to clinically defined AD-type dementia and 7 to non-AD dementias (See **Extended-Data Table-3** for dementia type). Cox proportional hazard models, adjusted for age, sex, education and cohort, demonstrated an increased risk for future progression to all-cause dementia in the A+T_NEO_+ (HR=39.9[15.6.-101.9], p<0.001) and A+T_MTL_+ (HR=5.6[1.3-24.2], p=0.02) groups compared to the A-T-(reference) group. There was no difference between the A+T- and the A-T-group (HR=1.5[0.4-5.2], p=0.53). Additionally, the A+T_NEO_+ group differed from both the A+T_MTL_+ (p=0.01) and the A+T-(p<0.001) groups, but the difference between A+T_MTL_+ and A+T-groups did not reach significance (p=0.09). Similar results were found when using progression to AD-type dementia (11 A+T_NEO_+, 2 A+T_MTL_+ and 1 A+T-) as outcome instead of all-cause dementia (**Extended-Data Fig.2**).

### Cognitive trajectories

Linear mixed effect models adjusting for age, sex, education and cohort indicated that the A+T_NEO_+ (standardized β [stβ] of interaction with time in months ± standard error=-0.020±0.002, T=-10.14, p<0.001), A+T_MTL_+ (stβ=-0.017±0.002, T=-8.84, p<0.001) and A+T-(stβ=-0.005±0.001, T=-5.26, p<0.001) groups showed faster decline over time on the modified preclinical Alzheimer cognitive composite 5 [mPACC5^15^] compared to the A-T-(reference) group (**Fig.1f**). Additionally, the A+T_NEO_+ (T=-7.53, p<0.001) and A+T_MTL_+ (T=-6.21, p<0.001) groups progressed faster than the A+T-group but there was no difference between the A+T_NEO_+ and A+T_MTL_+ groups (T=-1.13, p=0.26). Exploratory study of the mPACC5 subcomponents showed that A+T_NEO_+ and A+T_MTL_+ groups did not differ on delayed episodic memory (stβ=-0.002±0.003, p=0.47), but the A+T_NEO_+ group showed faster decline on executive function (stβ=-0.007±0.002, p=0.003) and semantic memory (stβ=-0.009±0.003, p=0.007, **Extended-Data Fig.3**).

On the MMSE, the A+T_NEO_+ (β=-0.056±0.005, T=-11.55, p<0.001), A+T_MTL_+ (β=-0.024±0.005, T=-4.72, p<0.001) and A+T-(β=-0.008±0.002, T=-3.46, p<0.001) groups showed faster decline over time compared to the A-T-(reference) group (**Fig.1g**). The A+T_NEO_+ (T=-9.51, p<0.001) and A+T_MTL_+ (T=-3.04, p=0.002) groups progressed faster than the A+T-group, and the A+T_NEO_+ group declined faster than the A+T_MTL_+ group (T=-4.82, p<0.001). Cognitive trajectories on MMSE and mPACC5 for each cohort are displayed in **Extended-Data Fig.4**.

## DISCUSSION

To examine whether amyloid and tau PET positive cognitively unimpaired individuals are destined to decline, we performed a multi-center study in 1325 participants with on average ∼3.5 years of clinical follow-up data available. We found that A+T_NEO_+ and A+T_MTL_+ cognitively unimpaired individuals had clearly increased risk for future development of MCI and all-cause dementia, and showed steep trajectories of cognitive decline. Hence, evidence of advanced AD pathology provided by amyloid and tau PET is strongly associated with short-term clinical progression in initially cognitively unimpaired individuals. This supports the NIA-AA criteria-based classification of A+T+ cognitively unimpaired individuals as “preclinical AD”, especially when “T” is defined by PET. To consider A+T+ merely as risk factor, and not manifest disease, may be an underestimation of its malignancy.

Although the A+T_NEO_+ group was at increased risk for progression to all-cause dementia compared to the A+T_MTL_+ group, there were no differences in risk for progression to MCI. A potential explanation is that tau pathological changes in the MTL can cause severe enough memory loss leading to an individual being classified as MCI (but not dementia), while widespread tau pathology into the neocortex might be needed to produce a dementia syndrome. Supporting this hypothesis, we found that the A+T_NEO_+ group exhibited faster decline in global cognition, semantic fluency and executive function, but not in memory function, compared to the A+T_MTL_+ group (**Fig.1g, Extended Data Table-3**).

This study has several limitations. First, there are inherent challenges due to the multi-center study design, such as data harmonization and pooling, different amyloid and tau PET tracers, dissimilarities in acquisition protocols and analytical methods and use of different neuropsychological tests to generate mPACC5 scores. Relatedly, the varying rates of clinical progression in A+T_NEO_+ and A+T_MTL_+ groups across cohorts (**Extended Data Table-1**) could be due to chance given the relatively small number of T+ participants but might also be a result of different ascertainment and recruitment methods. Second, we may have underestimated the actual risk of A+T_NEO_+ and A+T_MTL_+ cognitively unimpaired individuals due to “consent” or “volunteer bias” (lower study participation among individuals at risk) and “informative censoring” (the tendency of people to drop-out when experiencing onset or worsening of symptoms).^16^ Third, we acknowledge that our design only allowed establishing relative risk and not lifetime risk, and we did not control for the competing risk of death in our survival analyses.

Future studies should test whether our findings are generalizable to more diverse populations in terms of ethnicity, socio-economic status and medical comorbidities. Furthermore, studies with longer follow-up duration and larger numbers of A+T_NEO_+ and A+T_MTL_+ cognitively unimpaired individuals will help refine the current findings.

## Data Availability

Due to the multicentric design of the study, access to individual participant data from each cohort would have to be made available through the PIs of the respective cohorts. Generally, anonymised data can be shared by request from qualified academic investigators for the purpose of replicating procedures and results presented in the article, as long as data transfer is in agreement with the data protection regulation at the institution and is approved by the local Ethics Review Board. Requests for data from the open-access part of HABS can be submitted to: https://habs.mgh.harvard.edu.

## CONTRIBUTORS

RO, CRJ and OH designed the study. RO, APB and CG had full access to raw data and carried out the final statistical analyses. RO, ABP, CRJ and OH wrote the manuscript and had the final responsibility to submit for publication. All other authors contributed demographic, clinical, biomarker and neuroimaging data, contributed to the interpretation of the results and critically reviewed the manuscript.

## DECLARATION OF INTERESTS

OH has acquired research support (for the institution) from ADx, AVID Radiopharmaceuticals, Biogen, Eli Lilly, Eisai, Fujirebio, GE Healthcare, Pfizer, and Roche. In the past 2 years, he has received consultancy/speaker fees from AC Immune, Amylyx, Alzpath, BioArctic, Biogen, Cerveau, Fujirebio, Genentech, Novartis, Roche, and Siemens. RO has given a lecture in a symposium sponsored by GE Healthcare (fee paid to the institution). S.P. has served on scientific advisory boards and/or given lectures in symposia sponsored by Biogen, Eli Lilly, Geras Solutions, and Roche. RCP has consulted for Roche, Merck, Biogen, Eisai, Genentech and Nestle and receives research funding from the NIH. WF has performed contract research for Biogen MA Inc, and Boehringer Ingelheim, has been an invited speaker at Boehringer Ingelheim, Biogen MA Inc, Danone, Eisai, WebMD Neurology (Medscape), Springer Healthcare, is consultant to Oxford Health Policy Forum CIC, Roche, and Biogen MA Inc., participated in advisory boards of Biogen MA Inc and Roche, and is a member of the steering committee of PAVE and Think Brain Health (all funding/fees were paid to the institution).

## CODE AVAILABILITY

The codes used for data analyses in our study can be requested from the corresponding authors (RO, OH).

## ACKNOWLEDGEMENTS

The authors would like to thank Prof. Bruno Dubois for advice on the nomenclature used for the IWG criteria in **Fig.1a**.

Data used in the preparation of this article were obtained from the Harvard Aging Brain Study (HABS - P01AG036694; https://habs.mgh.harvard.edu). The HABS study was launched in 2010, funded by the National Institute on Aging. and is led by principal investigators Reisa A. Sperling MD and Keith A. Johnson MD at Massachusetts General Hospital/Harvard Medical School in Boston, MA.

Research of Alzheimer center Amsterdam is part of the neurodegeneration research program of Amsterdam Neuroscience. Alzheimer Center Amsterdam is supported by Stichting Alzheimer Nederland and Stichting VUmc fonds. The chair of Wiesje van der Flier is supported by the Pasman stichting. The SCIENCe project receives funding from Gieskes-Strijbis fonds and stichting Dioraphte.

## FUNDING

This project has received funding from the European Research Council (ERC) under the European Union’s Horizon 2020 research and innovation programme (grant agreement No. 949570, PI: Rik Ossenkoppele).

Work at Lund University was supported by the Swedish Research Council (2016-00906), the Knut and Alice Wallenberg foundation (2017-0383), the Marianne and Marcus Wallenberg foundation (2015.0125), the Strategic Research Area MultiPark (Multidisciplinary Research in Parkinson’s disease) at Lund University, the Swedish Alzheimer Foundation (AF-939932), the Swedish Brain Foundation (FO2021-0293), The Parkinson foundation of Sweden (1280/20), the Konung Gustaf V:s och Drottning Victorias Frimurarestiftelse, the Skåne University Hospital Foundation (2020-O000028), Regionalt Forskningsstöd (2020-0314) and the Swedish federal government under the ALF agreement (2018-Projekt0279). Acknowledgement is also made to the donors of the Alzheimer’s Disease Research, a program of the BrightFocus Foundation, for support of this research (A2021013F). APB is supported by a postdoctoral fellowship from the Fonds de recherche en Santé Québec (298314). The precursor of ^18^F-flortaucipir was provided by AVID radiopharmaceuticals and the precursor of ^18^F-RO948 was provided by Roche. The precursor of ^18^F-flutemetamol was sponsored by GE Healthcare. The MCSA is supported by the National Institute on Aging, U01 AG006786, GHR and the Alzheimer’s Association.

Research of Alzheimer center Amsterdam has been funded by ZonMW, NWO, EU-FP7, EU-JPND, Alzheimer Nederland, Hersenstichting CardioVascular Onderzoek Nederland, Health∼Holland, Topsector Life Sciences & Health, stichting Dioraphte, Gieskes-Strijbis fonds, stichting Equilibrio, Edwin Bouw fonds, Pasman stichting, stichting Alzheimer & Neuropsychiatrie Foundation, Philips, Biogen MA Inc, Novartis-NL, Life-MI, AVID, Roche BV, Fujifilm, Combinostics. WF is recipient of ABOARD, which is a public-private partnership receiving funding from ZonMW (#73305095007) and Health∼Holland, Topsector Life Sciences & Health (PPP-allowance; #LSHM20106).

The funding sources had no role in the design and conduct of the study; in the collection, analysis, interpretation of the data; or in the preparation, review, or approval of the manuscript.

## METHODS

### Participants

We included 1325 participants from the Mayo Clinic Olmsted Study of Aging^17^ (MCSA, n=680), the Swedish BioFINDER-1 (n=56) and BioFINDER-2 (n=228) studies at Lund University^7,10^, the Berkeley Aging Cohort study^18^ (BACS, n=109), the Harvard Aging Brain Study^19^ (HABS, n=162, data obtained in March 2022 from data release 2.0 via https://habs.mgh.harvard.edu), the Australian Imaging Biomarkers and Lifestyle Study of Ageing^20^ (AIBL, n=48) and the SCIENCe project^21^, which is part of the Amsterdam Dementia Cohort (ADC, n=42). A brief description of each cohort is provided in **Extended-Data Table-4**. All participants were i) cognitively unimpaired at baseline defined by neuropsychological test scores within the normative range given an individuals’ age, sex and educational background, ii) had amyloid PET available to determine Aβ status, iii) underwent a tau PET scan before January 1, 2019, to allow for sufficiently long follow-up duration, and iv) had at least one clinical follow-up visit available. Follow-up data was collected until April 1^st^, 2022. Written informed consent was obtained from all participants and local institutional review boards for human research approved the study.

### Amyloid PET status

Aβ status was determined using center-specific cut-offs or visual read metrics using [^18^F]flutemetamol PET for BioFINDER-1 and BioFINDER-2, [^11^C]Pittsburgh compound-B PET for MCSA, BACS and HABS, [^18^F]florbetapir PET for ADC and AIBL (n=47/48) and [^18^F]NAV4694 for AIBL (n=1/48), see **Extended-Data Table-5** for details.

### Tau PET status

Tau PET was performed using [^18^F]flortaucipir across all cohorts, except BioFINDER-2 where [^18^F]RO948 was used, and data were processed following previously described procedures (see **Extended-Data Table-6**). We computed tau PET status for a medial temporal lobe (MTL; unweighted average of bilateral entorhinal cortex and amygdala) and a neocortical (NEO; weighted average of bilateral middle temporal and inferior temporal gyri) region-of-interest.^11,22^ The threshold was determined for each cohort separately, based on the mean +2*standard deviation across all ββ-negative participants within each cohort (see cohort-specific cut-offs in **Extended-Data Table-6**). Based on amyloid and tau PET status we generated four different biomarker groups: A-T-, A+T-, A+T_MTL_+ (defined as tau PET positive in the MTL but not in the neocortex) and A+T_NEO_+ (defined as tau PET positive in the neocortex and/or in the MTL; 49/65 were also T_MTL_+). The A-T+ group was considerably smaller than the other groups (n=34, **Extended-Data Table-2**), hence their results are only reported in **Extended-Data Fig.1**.

### Clinical outcome measures

We used both binary and continuous measures of clinical progression. First, we examined progression from cognitively unimpaired to MCI (**Fig.1b**), all-cause dementia (**Fig.1c**) or AD-type dementia (**Extended Data Fig.2**). MCI was established using the Petersen criteria^23^ and is defined as significant cognitive symptoms as assessed by a physician, in combination with cognitive impairment on one or multiple domains (e.g., memory, executive functioning, attention, language) that is below the normative range given an individuals’ age, sex and educational background, but not sufficiently severe to meet diagnostic criteria for dementia. A large systematic review assessing 11,000 studies showed convergence across practices when validated diagnostic tools were used, such as in the current study.^24^ AD-type dementia was diagnosed using established criteria.^25^ Both MCI and dementia diagnoses were made by clinicians who were blinded for any PET or CSF outcome. For BACS, no formal diagnosis of MCI or dementia is made during the study, hence the cohort was excluded from this analysis. Second, we examined cognitive trajectories using a sensitive composite measure specifically developed to detect cognitive changes in preclinical stages of AD (i.e., the modified preclinical Alzheimer cognitive composite 5 [mPACC5^15,26^], **Fig.1f**) and a screening tests of global cognition (i.e., the Mini-Mental State Examination [MMSE] that is frequently used in clinical practice and in trials, **Fig.1g**). The mPACC5 consists of tests capturing episodic memory, executive function, semantic memory and global cognition.^15^ Individual tests were z-transformed using the baseline test scores of ββ-negative participants in each cohort as reference group and then averaged to obtain a composite z-score. The composition of mPACC5 is described for each cohort in **Extended-Data Table-7**.

### Statistical analyses

All statistical analyses were performed in R version 4.0.5. Differences in baseline characteristics between groups were assessed using ANOVA with *post hoc* T-tests with Bonferroni correction for continuous variables and χ^2^ and Kruskal-Wallis with *post hoc* Mann-Whitney U-tests for categorical or ordinal variables. First, we examined progression from cognitively unimpaired to MCI (**Fig.1b**), all-cause dementia (**Fig.1c**) or AD-type dementia (**Extended Data Fig.2**) using Cox proportional hazard models, adjusting for age, sex, education and cohort using A-T-as the reference group. To compare the other groups, we performed pairwise log-rank tests with a false discovery rate (FDR) correction. For individuals who progressed to MCI and subsequent dementia we used the respective times at conversion to MCI and to dementia for the analyses presented in **Fig.1b-c**. Second, we examined differences in cognitive trajectories between groups on the mPACC5 (**Fig.1f**) and on global cognition (i.e., MMSE, **Fig.1g**) using linear mixed effect models with random intercepts and slopes, adjusting for age, sex, education and cohort. Statistical significance for all models was set at p<0.05 two-sided.

## Notes

### Competing Interest Statement

The authors have declared no competing interest.

### Funding Statement

This project has received funding from the European Research Council (ERC) under the European Unions Horizon 2020 research and innovation programme (grant agreement No. 949570, PI: Rik Ossenkoppele).
Work at Lund University was supported by the Swedish Research Council (2016-00906), the Knut and Alice Wallenberg foundation (2017-0383), the Marianne and Marcus Wallenberg foundation (2015.0125), the Strategic Research Area MultiPark (Multidisciplinary Research in Parkinsons disease) at Lund University, the Swedish Alzheimer Foundation (AF-939932), the Swedish Brain Foundation (FO2021-0293), The Parkinson foundation of Sweden (1280/20), the Konung Gustaf V och Drottning Victorias Frimurarestiftelse, the Skane University Hospital Foundation (2020-O000028), Regionalt Forskningsstod (2020-0314) and the Swedish federal government under the ALF agreement (2018-Projekt0279). Acknowledgement is also made to the donors of the Alzheimers Disease Research, a program of the BrightFocus Foundation, for support of this research (A2021013F). APB is supported by a postdoctoral fellowship from the Fonds de recherche en Sante Quebec (298314). The precursor of 18F-flortaucipir was provided by AVID radiopharmaceuticals and the precursor of 18F-RO948 was provided by Roche. The precursor of 18F-flutemetamol was sponsored by GE Healthcare. The MCSA is supported by the National Institute on Aging, U01 AG006786, GHR and the Alzheimers Association.
Research of Alzheimer center Amsterdam has been funded by ZonMW, NWO, EU-FP7, EU-JPND, Alzheimer Nederland, Hersenstichting CardioVascular Onderzoek Nederland, Health~Holland, Topsector Life Sciences & Health, stichting Dioraphte, Gieskes-Strijbis fonds, stichting Equilibrio, Edwin Bouw fonds, Pasman stichting, stichting Alzheimer and Neuropsychiatrie Foundation, Philips, Biogen MA Inc, Novartis-NL, Life-MI, AVID, Roche BV, Fujifilm, Combinostics. WF is recipient of ABOARD, which is a public-private partnership receiving funding from ZonMW (#73305095007) and Health~Holland, Topsector Life Sciences & Health (PPP-allowance; #LSHM20106).
The funding sources had no role in the design and conduct of the study; in the collection, analysis, interpretation of the data; or in the preparation, review, or approval of the manuscript.

### Author Declarations

Written informed consent was obtained from all participants. For MSCA, this study was approved by institutional review boards at the Mayo Clinic and Olmsted Medical Center (both Rochester, MS, USA). For the BioFINDER-1 and BioFINDER-2 studies, ethical approval was given by the Regional Ethical Committee in Lund, Sweden. For BACS, the Institutional Review Board at Lawrence Berkeley National Laboratory and the University of California Berkeley approved the study. The AIBL study was approved by institutional ethics committees of Austin Health, St. Vincent's health, Hollywood Private Hospital and Edith Cowan University. For HABS, the protocol was approved by the Partners Human Research Committee. For ADC, the study protocol was approved by the Medical Ethics Review Committee of the Amsterdam UMC, location VU Medical center

