## Extended Data for "Amyloid and Tau PET positive cognitively unimpaired individuals: Destined to decline?"

**EXTENDED DATA CONTENT**

| **Table/Figure** | **Title** | **Page** |
| --- | --- | --- |
| Table 1 | Participant characteristics by cohort | 2 |
| Table 2 | Participant characteristics of the A-T+ group | 9 |
| Table 3 | Progression to non-AD dementia types and their AT status | 10 |
| Figure 1 | Main analyses now including the A-T+ group. | 11 |
| Figure 2 | Progression to AD dementia in the different categories AT categories | 12 |
| Figure 3 | Cognitive decline on the different mPACC5 components in the AT categories | 13 |
| Figure 4 | Cognitive decline on mPACC5 and MMSE in the individual cohorts | 14 |
| Table 4 | Cohorts descriptions | 15 |
| Table 5 | Methods to determine Amyloid PET status by cohort | 17 |
| Table 6 | Methods to determine Tau PET status by cohort | 18 |
| Table 7 | Composition of the mPACC5 for each cohort | 19 |
|  | Extended data - References | 20 |

**Extended Data Table 1.** Participant characteristics by cohort

**COHORT 1:** Mayo Clinic Olmsted Study of Aging (MCSA), n=665

|  | **A-T-** | **A+T-** | **A+T_MTL_+** | **A+T_NEO_+** |
| --- | --- | --- | --- | --- |
| N | 446 (67%) | 173 (26%) | 25 (4%) | 21 (3%) |
| Age, years | 66.8±9.2 | 76.4±8.3 | 75.1±6.7 | 79.5±6.3 |
| Sex, n (%) female | 209 (47%) | 80 (46%) | 12 (48%) | 6 (29%) |
| Education, years | 15.2±2.4 | 14.7±2.5 | 15.5±2.7 | 14.7±3.5 |
| MMSE, baseline score | 28.9±1.0 | 28.4±1.2 | 28.5±1.4 | 28.4±1.0 |
| Follow-up duration, months | 52.7±17.7 | 46.2±18.5 | 46.7±18.9 | 40.9±18.9 |
| Follow-up visits, number | 5.0±1.2 | 4.7±1.3 | 4.8±1.2 | 4.5±1.2 |
| Progression to MCI (%) | 21 (5%) | 16 (9%) | 11 (44%) | 6 (29%) |
| Progression to all-cause dementia (%) | 2 (0.4%) | 2 (1%) | 1 (4%) | 1 (5%) |

**COHORT 2:** BioFINDER-1, n=56

|  | **A-T-** | **A+T-** | **A+T_MTL_+** | **A+T_NEO_+** |
| --- | --- | --- | --- | --- |
| N | 29 (52%) | 15 (28%) | 5 (9%) | 7 (13%) |
| Age, years | 74.2±7.3) | 74.7±7.4 | 75.6±7.1 | 72.6±8.9 |
| Sex, n (%) female | 13 (45%) | 9 (60%) | 4 (80%) | 3 (43%) |
| Education, years | 12.6±3.9 | 12.1±3.9 | 9.2±1.5 | 12.1±2.1 |
| MMSE, baseline score | 29.0±1.1 | 29.7±0.5 | 29.0±0.7 | 27.6±1.6 |
| Follow-up duration, months | 37.6±8.4 | 34.1±9.0 | 36.0±5.3 | 36.0±10.0 |
| Follow-up visits, number | 2.4±0.7 | 2.4±0.6 | 3.0±0.0 | 3.1±1.2 |
| Progression to MCI (%) | 0 (0%) | 2 (13%) | 1 (20%) | 6 (86%) |
| Progression to all-cause dementia (%) | 0 (0%) | 1 (7%) | 0 (0%) | 4 (57%) |

**COHORT 3:** BioFINDER-2, n=222

|  | **A-T-** | **A+T-** | **A+T_MTL_+** | **A+T_NEO_+** |
| --- | --- | --- | --- | --- |
| N | 148 (67%) | 51 (23%) | 10 (5%) | 13 (6%) |
| Age, years | 65.3±10.1 | 70.1±9.3 | 74.1±6.8 | 75.8±7.7 |
| Sex, n (%) female | 75 (51%) | 23 (45%) | 6 (60%) | 9 (69%) |
| Education, years | 12.6±3.3 | 12.6.±3.8 | 11.2±4.0 | 11.1±3.5 |
| MMSE, baseline score | 29.0±1.2 | 28.8±1.3 | 28.0±1.6 | 28.7±1.0 |
| Follow-up duration, months | 29.1±10.7 | 36.3±8.8 | 39.4±5.1 | 37.1±9.8 |
| Follow-up visits, number | 2.5±0.8 | 3.5±1.0 | 4.1±0.7 | 3.2±1.0 |
| Progression to MCI (%) | 3 (2%) | 4 (8%) | 7 (70%) | 8 (62%) |
| Progression to all-cause dementia (%) | 1 (1%) | 0 (0%) | 1 (10%) | 4 (31%) |

**COHORT 4:** the Berkeley Aging Cohort study (BACS), n=107

|  | **A-T-** | **A+T-** | **A+T_MTL_+** | **A+T_NEO_+** |
| --- | --- | --- | --- | --- |
| N | 62 (58%) | 36 (34%) | 4 (4%) | 5 (5%) |
| Age, years | 77.1±7.1 | 76.7±4.4 | 81.0±2.3 | 75.8±3.0 |
| Sex, n (%) female | 36 (58%) | 23 (64%) | 1 (25%) | 3 (60%) |
| Education, years | 17.4±2.9 | 16.7±1.9 | 16.2±1.3 | 16.2±0.8 |
| MMSE, baseline score | 28.8±1.1 | 28.6±1.5 | 27.5±1.3 | 28.8±0.8 |
| Follow-up duration, months | 32.2±15.7 | 33.6±15.2 | 21.6±11.4 | 38.0±12.4 |
| Follow-up visits, number | 3.5±1.5 | 3.6±1.4 | 2.8±0.5 | 3.2±1.6 |
| Progression to MCI (%) | NA | NA | NA | NA |
| Progression to all-cause dementia (%) | NA | NA | NA | NA |

**COHORT 5:** the Harvard Aging Brain Study (HABS), n=155

|  | **A-T-** | **A+T-** | **A+T_MTL_+** | **A+T_NEO_+** |
| --- | --- | --- | --- | --- |
| N | 103 (66%) | 38 (25%) | 5 (3%) | 9 (6%) |
| Age, years | 75.0±6.3 | 77.3± 6.3 | 76.8±5.7 | 77.4±5.3 |
| Sex, n (%) male | 56 (54%) | 21 (55%) | 4 (80%) | 7 (78%) |
| Education, years | 15.9±3.3 | 16.1±3.0 | 16.4±2.6 | 17.3±1.4 |
| MMSE, baseline score | 29.3±1.0 | 29.2±0.9 | 29.0±1.4 | 28.3±1.2 |
| Follow-up duration, months | 29.2±12.6 | 26.2±14.8 | 23.1±14.1 | 21.9±4.6 |
| Follow-up visits, number | 3.6±1.0 | 3.4±1.0 | 3.2±1.1 | 3.2±0.4 |
| Progression to MCI (%) | 1 (1%) | 3 (8%) | 2 (40%) | 3 (33%) |
| Progression to all-cause dementia (%) | 0 (0%) | 0 (0%) | 0 (0%) | 0 (0%) |

**COHORT 6:** the Australian Imaging Biomarkers and Lifestyle Study of Ageing (AIBL), n=46

|  | **A-T-** | **A+T-** | **A+T_MTL_+** | **A+T_NEO_+** |
| --- | --- | --- | --- | --- |
| N | 30 (65%) | 9 (20%) | 3 (7%) | 4 (9%) |
| Age, years | 72.2±5.5 | 78.9±8.0 | 84.7±0.6 | 76.5±6.6 |
| Sex, n (%) male | 17 (57%) | 6 (67%) | 2 (67%) | 3 (75%) |
| Education, years | 12.7±2.4 | 13.3±2.1 | 9.5±1.7 | 11.6±2.2 |
| MMSE, baseline score | 28.9±1.0 | 28.8±1.6 | 26.3±2.5 | 26.2±2.4 |
| Follow-up duration, months | 50.5±14.9 | 46.6±20.0 | 39.5±4.3 | 47.0±17.1 |
| Follow-up visits, number | 3.7±0.9 | 3.9±1.4 | 2.7±0.6 | 3.0±1.2 |
| Progression to MCI (%) | 1 (3%) | 1 (11%) | 2 (67%) | 4 (100%) |
| Progression to all-cause dementia (%) | 0 (0%) | 0 (0%) | 0 (0%) | 2 (50%) |

Education was originally recorded as categorical data, i.e., 7-8 years, 9-12 years, 13-15 years or 15+ years. To be able to include this variable with the other cohorts, we converted it to the average of each category, or to 15 for the last category.

**COHORT 7:** the Amsterdam Dementia Cohort (ADC), n=40

|  | **A-T-** | **A+T-** | **A+T_MTL_+** | **A+T_NEO_+** |
| --- | --- | --- | --- | --- |
| N | 25 (63%) | 6 (15%) | 3 (8%) | 6 (15%) |
| Age, years | 63.1±7.0 | 71.0±6.4 | 69.7±2.5 | 70.0±4.0 |
| Sex, n (%) male | 13 (52%) | 2 (33%) | 2 (67%) | 3 (50%) |
| Education, years | 12.5±2.6 | 12.5±3.8 | 10.0±0 | 13.3±3.1 |
| MMSE, baseline score | 28.9±1.1 | 28.2±1.5 | 28.7±1.2 | 28.0±1.4 |
| Follow-up duration, months | 33.4±9.6 | 27.5±9.6 | 27.0±8.5 | 33.3±12.2 |
| Follow-up visits, number | 4.1±0.8 | 3.8±0.8 | 3.3±0.6 | 3.7±0.5 |
| Progression to MCI (%) | 0 (0%) | 0 (0%) | 2 (67%) | 5 (83%) |
| Progression to all-cause dementia (%) | 1 (4%) | 0 (0%) | 0 (0%) | 1 (17%) |

**Extended Data Table 2.** Participant characteristics of the A-T+ group

|  | **A-T+** |
| --- | --- |
| N | 34 |
| Age, years | 76.4±9.3 |
| Sex, n (%) male | 19 (55.9) |
| Education, years | 14.9±3.0 |
| Follow-up duration, months | 38.1±20.2 |
| Follow-up visits, number | 3.9±1.4 |
| MMSE, baseline score | 28.7±1.0 |
| Cohort | MCSA = 15  BioFINDER-1 = 0  BioFINDER-2 = 6  BACS = 2  HABS = 7  AIBL = 2  ADC = 2 |

**Extended Data Table 3.** Progression to non-AD dementia types and their AT status

| **Type of Dementia** | **N** | **AT status** |
| --- | --- | --- |
| Dementia with Lewy bodies | 2 | A-T- (both) |
| Behavioral variant frontotemporal dementia | 1 | A+T- |
| Vascular dementia | 1 | A-T- |
| Progressive supranuclear palsy | 1 | A-T- |
| Parkinson’s disease dementia | 1 | A+T- |
| Aphasia due to cerebrovascular disease | 1 | A+T_NEO_+ |

**Extended Data Figure 1.** Main analyses from Figure 1, but now also including the A-T+ group.

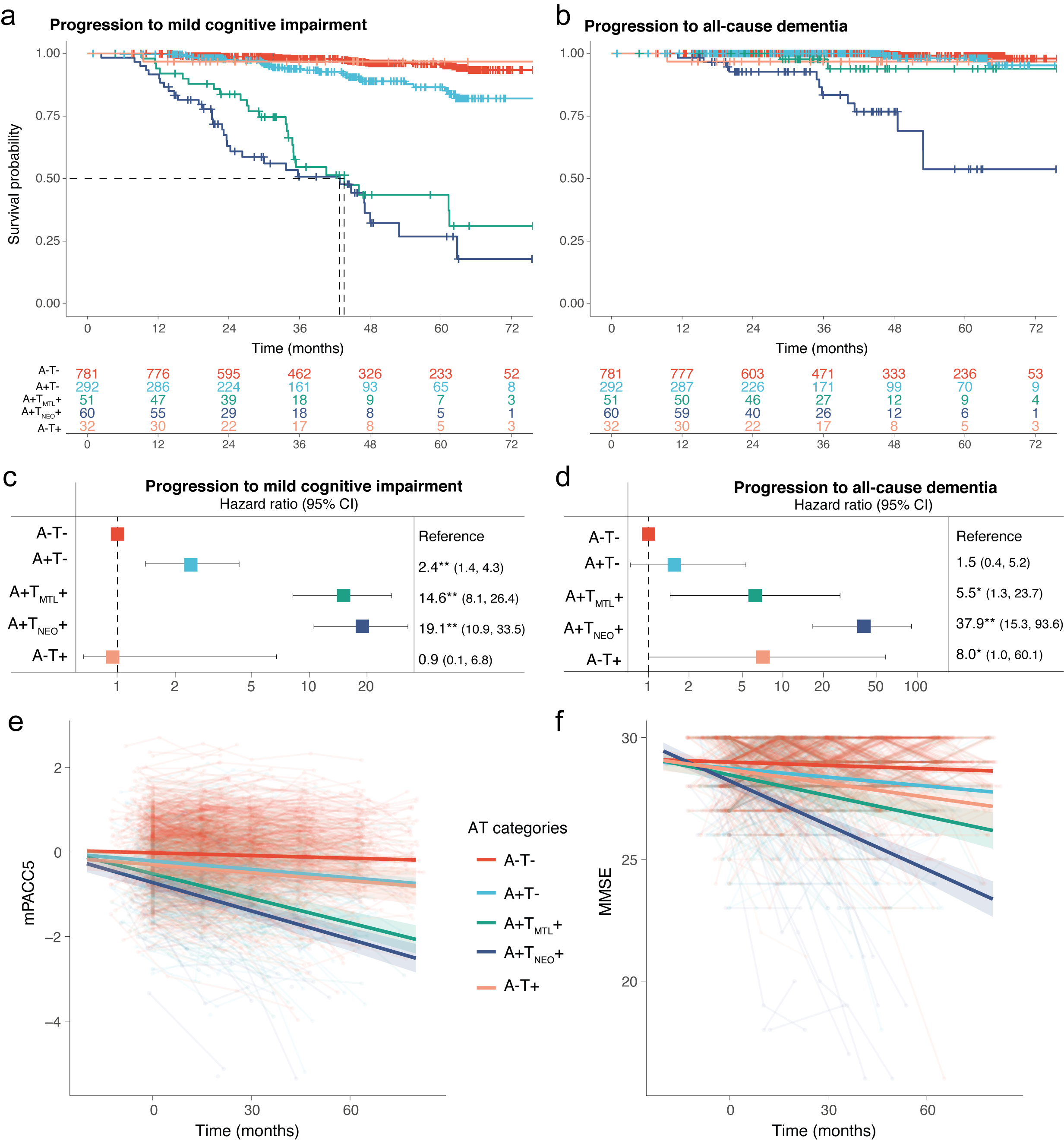

This figure resembles figure 1 of the main manuscript but now also includes the A-T+ group. **a**, Survival curves in relation to progression to MCI in the different AT categories, with the table of total number of participants available at each time point. **b**, Survival curves in relation to progression to all-cause dementia in the different AT categories, with the table of total number of participants available at each time point. **c, d** Forest plots showing the hazard ratios from the survival analyses shown in a and b, from Cox regression models including age, sex, education, and cohort as covariates. **e**, Cognitive trajectories of mPACC5 scores over time in the different AT categories **f**, Cognitive trajectories of MMSE scores over time in the different AT categories. The average regression line for each group was plotted from linear mixed effect models including age, sex, education, and cohort as covariates. Data are anchored to the tau-PET visit (Time 0), and cognitive data up to 1 year prior to PET was included. *p=0.01, **p<0.001.

**Extended Data Figure 2.** Progression to AD dementia in the different categories AT categories

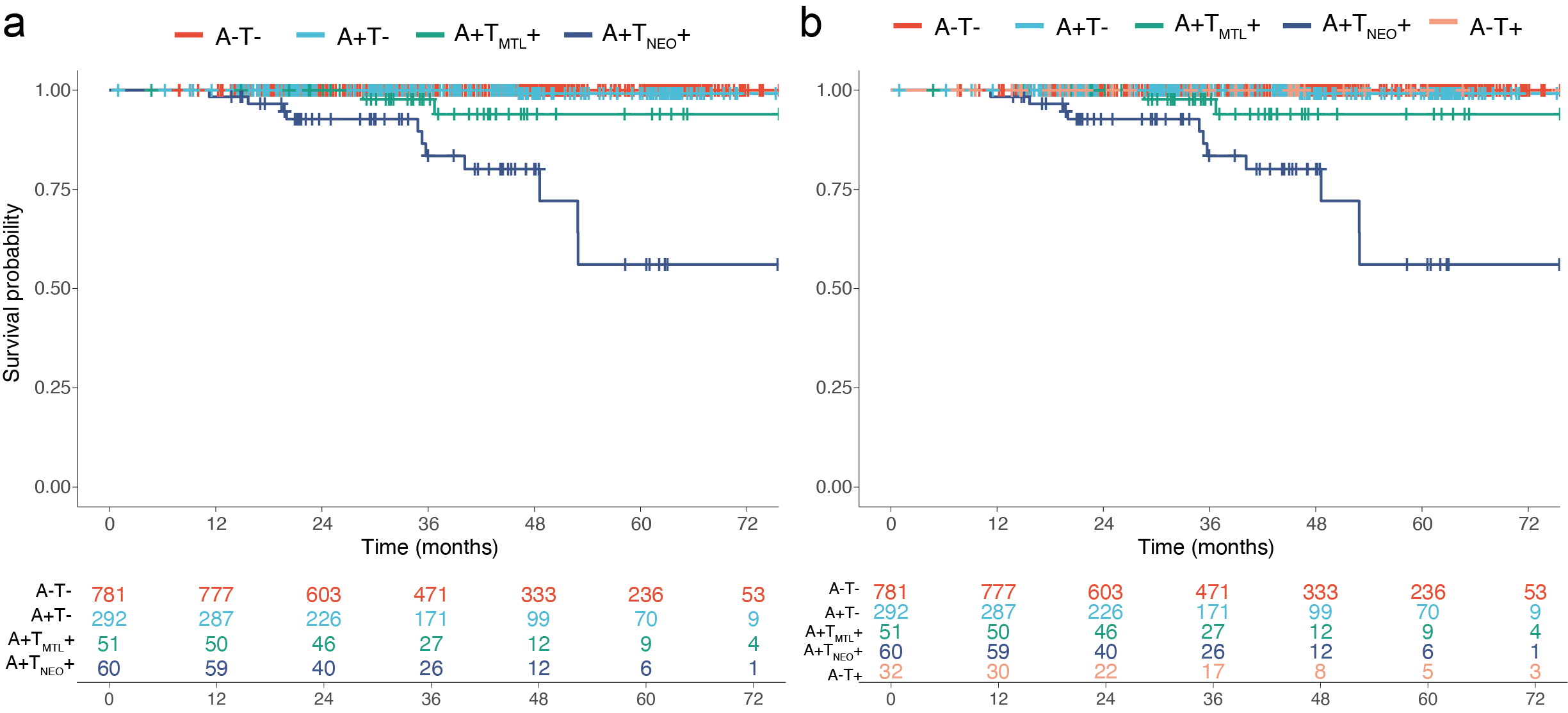

**a**, Survival curves in relation to progression to AD dementia in the main AT categories, with the table of total number of participants available at each time point. **b**, Survival curves in relation to progression to AD dementia when including the A-T+ group, with the table of total number of participants available at each time point.

Given the small number of events, the hazard ratios are difficult to interpret, but the A+T-, A+T_MTL_+ and A+T_NEO_+ groups had higher HR’s compared to A-T- reference group (all p<0.001). Pairwise comparisons indicated that both the A+T_NEO_+ (p<0.001) and A+T_MTL_+ (p=0.008) groups differed from the A+T- group, and the A+T_NEO_+ group differed from the A+T_MTL_+ group (p=0.01).

**Extended Data Figure 3.** Cognitive decline on the different mPACC5 components in the AT categories

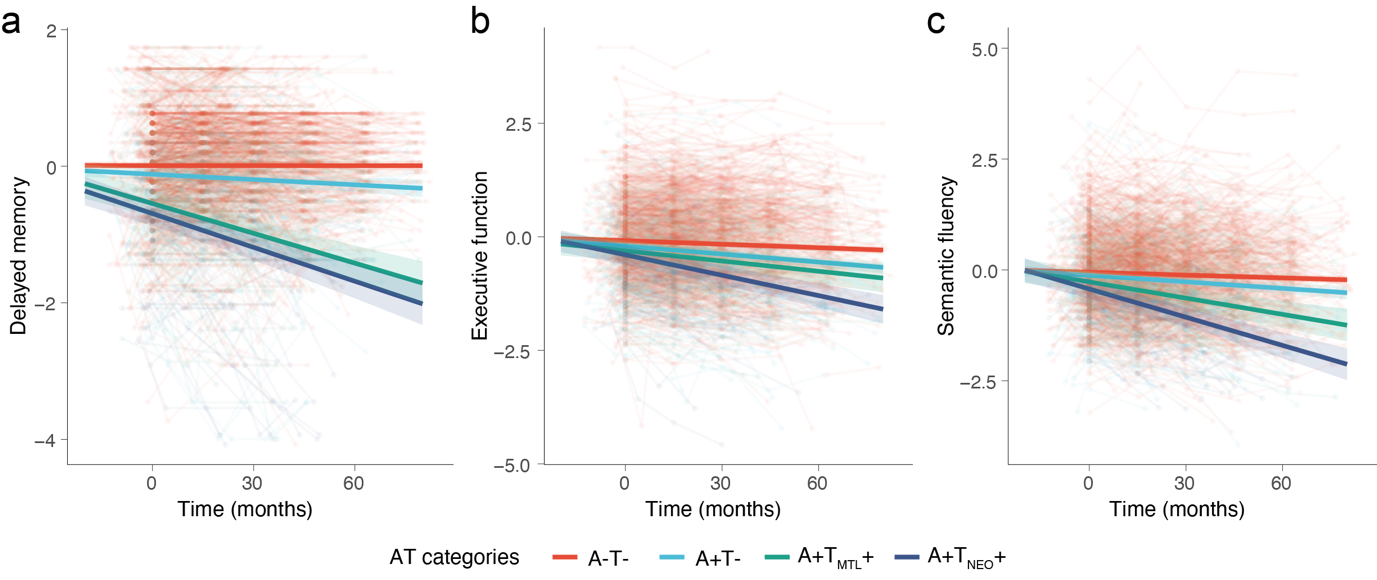

**a**, Cognitive trajectories on the delayed memory component of the mPACC5 over time in the different AT categories **b**, Cognitive trajectories on the executive function component of the mPACC5 over time in the different AT categories. **c**, Cognitive trajectories on the semantic fluency component of the mPACC5 over time in the different AT categories The average regression line for each group was plotted from linear mixed effect models including age, sex, education, and cohort as covariates. Data are anchored to the tau-PET visit (Time 0), and cognitive data up to 1 year prior to PET was included.

**Extended Data Fig 4.** Cognitive decline on mPACC5 and MMSE in the individual cohorts

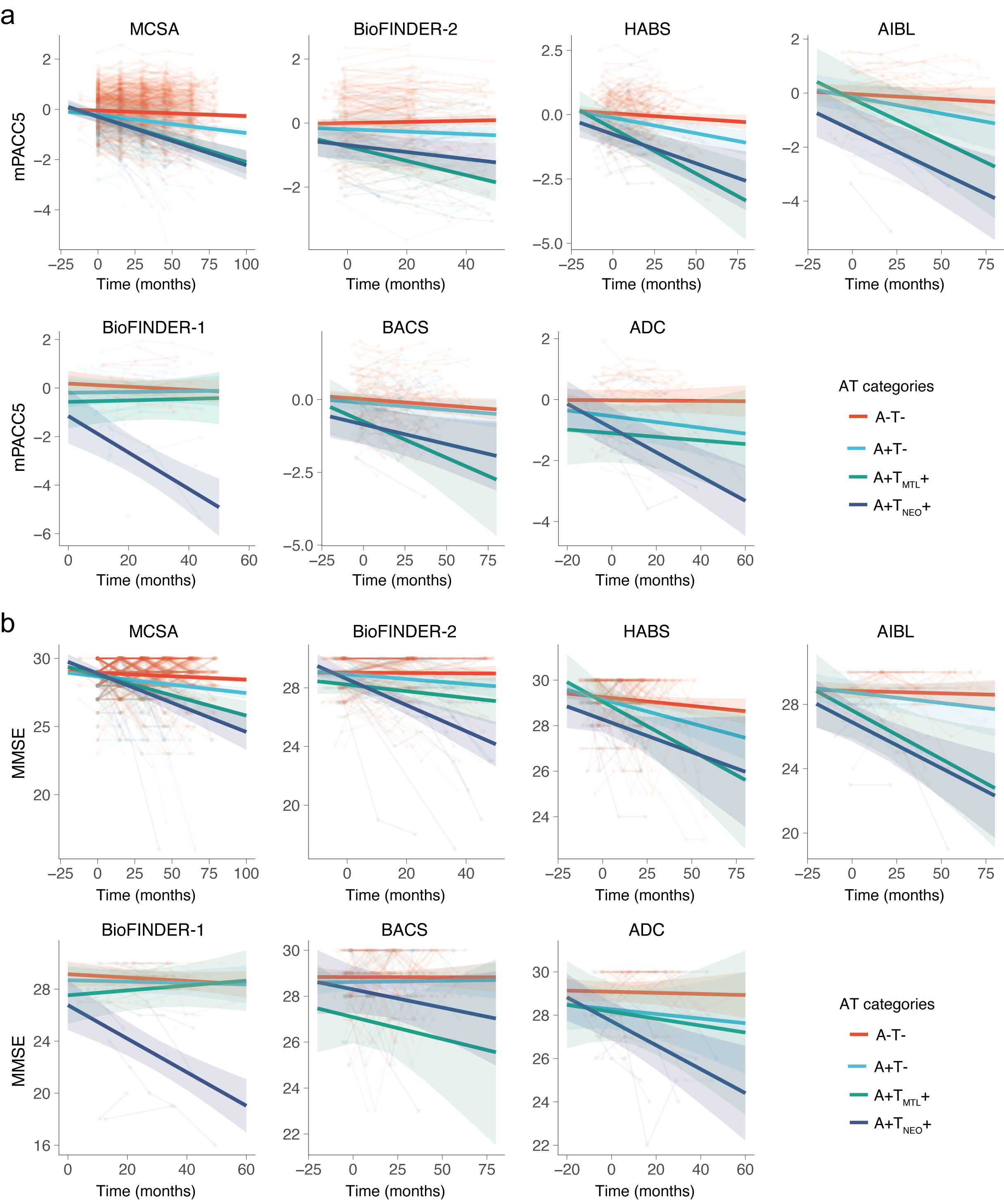

**a**, Cognitive trajectories of mPACC5 scores over time in the different AT categories in each individual cohort. **b**, Cognitive trajectories of MMSE scores over time in the different AT categories in each individual cohort. The average regression line for each group was plotted from linear mixed effect models including age, sex, and education as covariates. Data are anchored to the tau-PET visit (Time 0), and cognitive data up to 1 year prior to PET was included.

**Extended Data Table 4.** Cohort descriptions

| **Cohort** | **Cohort description** | **References** |
| --- | --- | --- |
| BioFINDER-1 &  BioFINDER-2 | The Swedish BioFINDER studies are longitudinal studies covering the entire AD continuum in which participants were recruited at Skåne University Hospital and the Hospital of Angelholm, Sweden. The main inclusion criteria were absence of cognitive symptoms as assessed by a physician with special interest in cognitive disorders, being fluent in Swedish, having no significant unstable systemic illness that made it difficult to participate in the study, having no current significant alcohol or substance misuse, and no significant neurological or psychiatric illness. For the current study participants above > 50 years old were included. Both cognitively healthy older adults and SCD participants were included. The SCD participants were referred from participating memory clinic because of cognitive complaints, but did not fulfill criteria for MCI (defined using criteria by Petersen and operationalized according to^1,2^) following a neuropsychological test battery. | ^3-5^ |
| MCSA | The Mayo Clinic Study of Aging (MCSA) is a longitudinal population-based study of cognitive aging in Olmsted County, Minnesota. The study was designed to study prevalence, incidence and risk factors for MCI and dementia. Potential participants are randlomly enumerated from the Olmsted County, MN, census and enrolled by age/sex strata. Enumeration is repeated to maintain a sample of approximately 3000 active participants. At entry, every person underwent evaluations that included a medical history review and interview with the participant and a study partner, a neurological examination by a physician; and a neuropsychological examination. For this study, participants were considered MCI only if the study coordinator, physician, and neuropsychologist were all in agreement regarding the MCI diagnosis. Participants were judged cognitively normal if they did not meet MCI criteria. Participants aged between 50 and 89 years old were included in the current study. | ^6^ |
| BACS | The Berkeley Aging Cohort Study (BACS) is a community-dwelling cohort that is a convenience sample of healthy individuals who are older than 60 years and reside in the San Francisco Bay Area of California. Inclusion criteria were no impairment of activities of daily living, absence of any neurological or psychiatric condition that potentially affects brain structure and function, no cognitive complaints, normal perform ance on cognitive tests (maximally 1.5 standard deviation below age-, education-, and sex-adjusted norms), no use of psychoactive drugs and absence of sensory impairment that might interfere with cognitive testing.  MCI or dementia diagnosis was not available in BACS and thus this cohort was included in analyses of cognitive trajectories only. | ^7^ |
| HABS | The Harvard Aging Brain Study (HABS) is a a longitudinal study on aging and AD, from Memory Disorders Clinics at the Massachusetts General and Brigham and Women’s Hospitals, and from the Massachusetts Alzheimer’s Disease Research Center. The cohort includes cognitively normal, healthy older individuals according to the following criteria. Inclusion criteria: 65 years of age or older, a CDR score of 0, MMSE>25, scores above age- and education-adjusted norms on the 30-Minute Delayed Recall of the Logical Memory Story A, and a score of less than 11 on the Geriatric Depression Scale. Exclusion criteria: history of alcoholism, drug abuse, head trauma, or current serious medical/psychiatric illness.  Data was obtained in March 2022 from data release 2.0 via [https://habs.mgh.harvard.edu](https://habs.mgh.harvard.edu/) | ^8^ |
| AIBL | The Australian Imaging, Biomarker & Lifestyle Flagship Study of Ageing (AIBL) is a longitudinal, prospective cohort with participants coming from two-site study – Melbourne and Perth. To be included in the study, participants were (1) ≥60 years old; (2) fluent in English; (4) had completed at least 7 years of education; (5) did not have any history of neurological or psychiatric disorders, drug or alcohol abuse or dependence, or any other unstable medical condition; and (6) were deemed to be cognitively unimpaired (CU), based on their performance on a battery of cognitive assessments that AIBL participants undergo every 12 to 18 months. A multidisciplinary clinical review panel determines whether an individual is CU, based on the available clinical and neuropsychological information. | ^9,10^ |
| ADC | The Amsterdam Dementia Cohort (ADC) is a prospective cohort study including patients with subjective cognitive decline (SCD) presenting at the Alzheimer Center of the VU University Medical Center Amsterdam. All participants have been referred to the memory clinic by their general practitioner, and a neurologist or geriatrician in the case of a second opinion for evaluation of cognitive complaints. They receive standardized dementia screening at the memory clinic, including an interview with a neurologist, physical and neurological examination, neuropsychological assessment. The main inclusion criteria were a diagnosis of SCD (i.e., cognitive complaints and normal cognition) and age ≥ 45 years. Exclusion criteria are MCI, dementia, major psychiatric disorder (i.e., current depression, personality disorders, schizophrenia), neurological diseases known to cause memory complaints (i.e., Parkinson’s disease, epilepsy), HIV, abuse of alcohol or other sub- stances, and language barrier. | ^11^ |

**Extended Data Table 5.** Methods to determine Amyloid PET status by cohort

| **Cohort** | **Tracer** | **Methodology** | **Cut-off** | **References** |
| --- | --- | --- | --- | --- |
| BioFINDER-1 | [^18^F]flutemetamol | Global neocortical composite standardized uptake value ratios (SUVR) for the 90-110min interval p.i. with whole cerebellum as reference region | >1.03 SUVR | ^12,13^ |
| BioFINDER-2 | [^18^F]flutemetamol | Global neocortical composite SUVR for the 90-110min interval p.i. with whole cerebellum as reference region | >1.03 SUVR | ^12,13^ |
| MCSA | [^11^C]PIB | Late uptake amyloid PET images were acquired from 40-60 minutes p.i. A meta-ROI was calculated as the voxel-number weighted average of uptake in a target region including prefrontal, orbitofrontal, parietal, temporal, anterior and posterior cingulate, and precuneus regions divided by the uptake in the cerebellar crus gray matter. | >1.48 SUVR  (>21CL) | ^14^ |
| BACS | [^11^C]PIB | Distribution volume ratio (DVR) images were calculated with Logan graphical analysis over 35–90 min data and normalized to a cerebellar gray reference region. | >1.065 DVR | ^15^ |
| HABS | [^11^C]PIB | Global neocortical composite DVR from a 60-minute dynamic acquisition p.i. with cerebellar gray matter as reference tissue. | >1.2 DVR (>26 CL) | ^16,17^ |
| AIBL | [^11^C]PIB/  [^18^F]NAV4694 | The standard Centiloid (CL) cortical and whole cerebellar volumes of interest template were applied to the summed and spatially normalised PET images in order to obtain SUVR’s. These SUVR were transformed into CL units by linear transformation using the PET tracer-specific equations published for conversion of CL method SUVR to CL units. | >24 CL | ^18^ |
| ADC | [^18^F]florbetapir | Visual read following guidelines provided by Avid Radiopharmaceuticals corresponding to >17 CL. | - | ^19,20^ |

CL = Centiloid; DVR = Distribution volume ratio; SUVR = Standardized uptake value ratio.

Centiloid (CL) units were presented when available.

**Extended Data Table 6.** Methods to determine Tau PET status in the medial temporal lobe (MTL) and neocortex (NEO) by cohort

| **Cohort** | **Tracer** | **Scanning interval** | **Reference region** | **Reference** | **Cut-off MTL** | **Cut-off NEO** |
| --- | --- | --- | --- | --- | --- | --- |
| BioFINDER-1 | [^18^F]flortaucipir | 80-100min p.i. | Inferior cerebellar GM | ^3^ | 1.26 SUVR | 1.29 SUVR |
| BioFINDER-2 | [^18^F]RO948 | 70-90min p.i. | Inferior cerebellar GM | ^5^ | 1.34 SUVR | 1.36 SUVR |
| MCSA | [^18^F]flortaucipir | 80-100min p.i. | Cerebellar crus GM | ^14^ | 1.30 SUVR | 1.37 SUVR |
| BACS | [^18^F]flortaucipir | 80-100min p.i. | Inferior cerebellar GM | ^21^ | 1.36 SUVR | 1.32 SUVR |
| HABS | [^18^F]flortaucipir | 80-100min p.i. | Cerebellar GM | ^16^ | 1.36 SUVR | 1.28 SUVR |
| AIBL | [^18^F]flortaucipir | 80-100min p.i. | Cerebellar GM | ^22^ | 1.31 SUVR | 1.38 SUVR |
| ADC | [^18^F]flortaucipir | 80-100min p.i. | Cerebellar GM | ^23^ | 1.26 SUVR | 1.26 SUVR |

GM = Gray matter; MTL = Medial temporal lobe; NEO = Neocortical; p.i. = Post-injection; SUVR = Standardized uptake value ratio.

The cut-offs were generated in each individual cohort, based on the mean + 2*standard deviation across all Aβ-negative participants within each cohort. We computed tau PET status for a medial temporal lobe (MTL; unweighted average of bilateral entorhinal cortex and amygdala) and a neocortical (NEO; weighted average of bilateral middle temporal and inferior temporal gyri) region-of-interest.

**Extended Data Table 7.** Composition of the mPACC5 for each cohort

| **Cohort** | **Global Cognition** | **Episodic Memory** | **Time executive function** | **Semantic memory** |
| --- | --- | --- | --- | --- |
| BioFINDER-1 | MMSE | ADAS-COG delayed word recall | Symbol digit modalities test | Animal fluency |
| BioFINDER-2 | MMSE | ADAS-COG delayed word recall | Symbol digit modalities test | Animal fluency |
| MCSA | MMSE^a^ | AVLT delayed recall | WAIS-R Digit Symbol | Sum of animal, fruits and vegetables fluency |
| BACS | MMSE | CVLT – Delayed recall | Symbol digit modalities test | Animal fluency |
| HABS | MMSE | SRT – Delayed recall | Symbol digit modalities test | Animal fluency |
| AIBL | MMSE | CVLT – Delayed recall | Symbol digit modalities test | Sum of animal and names fluency |
| ADC | MMSE | RAVLT – Delayed recall | TMT-B | Animal fluency |

Note that the episodic memory test was given double weight and thus accounted for 40% of the mPACC5 score.

^a^ A 38-point test, the Short Test of Mental Status (STMS)^24^, was converted to MMSE scores using an in-house developed algorithm^25^.
